# Acoustic and language analysis of speech for suicide ideation among US veterans

**DOI:** 10.1101/2020.07.08.20147504

**Authors:** Anas Belouali, Samir Gupta, Vaibhav Sourirajan, Jiawei Yu, Nathaniel Allen, Adil Alaoui, Mary Ann Dutton, Matthew J. Reinhard

## Abstract

U.S. veterans are 1.5 times more likely to die by suicide than Americans who never served in the military. Considering such high rates, there is an urgent need to develop innovative approaches for objective and clinically applicable assessments to detect individuals at high risk. We hypothesize that speech in suicidal veterans has a range of distinctive acoustic and linguistic features. The purpose of this work is to build an automated machine learning and natural language processing tool to screen for suicidality. Veterans made 588 narrative audio recordings via a mobile app in a real-life setting. In addition, veterans completed self-report psychiatric scales and questionnaires. Recordings were analyzed to extract voice characteristics including prosodic, phonation, and glottal. The audios were also transcribed to extract textual features for linguistic analysis. We evaluated the acoustic and linguistic features using both statistical significance and ensemble feature selection. We also examined the performance of different machine learning algorithms on multiple combinations of features to classify suicidal and non-suicidal audios. Random Forest classifier correctly identified suicidal ideation in veterans based on the combined set of acoustic and linguistic features of speech with 86% sensitivity, 70% specificity, and an area under the receiver operating characteristic curve (AUC) of 80%. Speech analysis of audios collected from veterans in everyday life settings using smartphones is a promising approach for suicidal ideation detection. A machine learning classifier may eventually help clinicians identify and monitor high-risk veterans.

## Introduction

Suicide prevention remains a challenging clinical issue, especially among Veterans. According to the most recent data of the United States (US) Veterans Affairs (VA), 17 veterans on average commit suicide per day and rates continue to rise.^1^ After controlling for factors like age and gender, Veterans faced a 1.5 times greater risk for suicide compared to civilians. From 2005 to 2017, the suicide rate in the US civilian population increased 22.4%, while rates among Veterans increased more than 49%.^1^ To help address such alarming rates, there is an urgent need to innovate and develop objective and clinically applicable assessments that could help detect high-risk individuals. Suicidal ideation is a known risk factor for suicide and has been found to be a predictor of immediate or long-term suicide attempts and deaths.^2,3^ Screening high-risk groups such as veterans for suicidal behavior is crucial for early detection and prevention.^4^

Currently, screening for suicide in a primary care setting is the result of a complex dynamic between provider and subject where the provider ultimately relies on the subject to disclose suicidal thoughts. To assess suicidality, healthcare providers use one of the several self-report screening tools such as the Suicidal Ideation Questionnaire (SIQ) or clinician-administered scales such as the Ask Suicide-Screening Questions (ASQ).^5,6^ Although shown to be sensitive in diagnosing suicidality, these types of testing require long visits to a clinician’s office and rely heavily on the honesty and disposition of a subject to communicate their symptoms. Additionally, implicit bias may affect the mental health assessment process and can result in misdiagnosis.^7^ Due to these limitations, research into finding objective markers to aid clinical assessment is key in the fight against suicide.

Recent advances in digital technologies and mHealth devices have the potential to provide novel data streams for suicide prevention research.^8^ Speech, for instance, is an information-rich signal and measurable behavior that can be collected outside the clinical setting, which can increase accessibility to care and enable real-time and context-aware monitoring of an individual’s mental state.^9,10^ In fact, several studies have investigated the characteristics of voice as an objective marker to understand various mental states and psychiatric disorders. Multiple research papers investigated voice in depression and many acoustic markers were identified.^9,11,12^ Recently, researchers were able to classify depressed and healthy speech using deep learning techniques applied to both audio and text features. ^13^ Another recent study investigated speech and PTSD in US veterans. The authors identified 18 acoustic features and built a classifier to differentiate the 54 PTSD veterans from 77 controls with an area under the ROC curve of 0.95.^14^ Using smartphones to collect voice data from 28 bipolar patients, one study performed classification of affective states (manic vs depression episodes) longitudinally based on voice features with accuracy in the range of 0.61–0.74.^15^ These studies and several others support the feasibility and validity of detecting different mental disorders from speech.

Studies analyzing the spoken language of suicidal patients date back as early as 1992, describing suicidal voices as sounding hollow, toneless, monotonous, with mechanical and repetitive phrasing, and a loss in intensity over an utterance.^9,16,17^ It has also been suggested that pre-suicidal mental state causes changes to speech production mechanisms which in turn alters the acoustic properties of speech in measurable ways.^17^ One study comparing suicidal and non-suicidal speech in 16 adolescents identified glottal features to show the strongest differences between the two groups. In particular, suicidal patients had lower Opening Quotient (OQ), and Normalised Amplitude Quotient (NAQ), acoustic measurements associated with more breathy voices.^18^ Acoustic features such as fundamental frequency (F0), amplitude modulation (AM), pauses and rhythm-based features were also investigated to differentiate between suicidal and depressed patients.^12,19^ More recent work used both linguistic and acoustic features of speech to classify 379 patients in one of three groups (suicidal, mentally ill but not suicidal, or controls) with accuracies in the range of 0.74-0.85.^20,21^ Although these are promising findings from a large sample, the authors didn’t explain which important acoustic and linguistic variables were selected in the models and if there were any significant acoustic features correlated with suicidality.

Our work investigates a machine learning (ML) approach using speech for the detection of suicidal ideation in US veterans. We hypothesize that speech in suicidal veterans has a range of distinctive acoustic and linguistic features that could identify suicide ideation in veterans. We investigate these features in 588 narrative audios collected longitudinally from 124 Veterans in a naturalistic setting using a mobile app that we developed for data collection purposes. We conduct comprehensive feature engineering on the recordings to extract several sets of features and then evaluate different classifiers and learning approaches. To our knowledge, this is the first study to investigate speech in suicidal veterans using a large number of audios collected in everyday life settings.

## Materials and Methods

### Study data and setting

Data for the present study was obtained as part of a larger intervention study for Gulf War Illnesses at the Washington DC VA Medical Center. 149 veterans meeting the Center for Disease Control’s criteria for Gulf War Illness^22^ were recruited for the study and of these, 124 participants submitted 588 recordings via an Android smartphone app developed for data collection. An Android tablet (Samsung Galaxy Tab 4) with the mobile app installed was provided to each veteran to enable study participation from home.

All data was collected longitudinally from veterans in a naturalistic setting using the smartphone app. At each time-point of the study (week 0, week 4, week 8, 3 months, 6 months, 1 year), participants received reminder notifications and were prompted to complete multiple assessments which included several self-report psychiatric scales and questionnaires. Veterans responded via audio recordings to open-ended questions about their general health in the past weeks/months and about their expectations from the study. These audio recordings were gathered for potential future qualitative analysis.

Each recording response had a Patient Health Questionnaire (PHQ-9) administered as part of the health questionnaire battery. Item-9 of the PHQ-9^23^ is commonly used in research to screen for suicidality and has been validated to be predictive of suicide in both the general population and in US veterans.^24,25^ It asks, “Over the last two weeks, how often have you been bothered by thoughts that you would be better off dead or of hurting yourself in some way?” Response options are “not at all”, “several days”, “more than half the days”, or “nearly every day”. We considered a subject as suicidal at the time of recording, if they answered with any option other than “not at all”.

### Feature extraction and preprocessing

Voice features can be divided into acoustic and linguistic features. We conducted comprehensive feature engineering on each recording to extract several sets of features. The study procedure is detailed in **Figure 1**.

**Figure 1:**
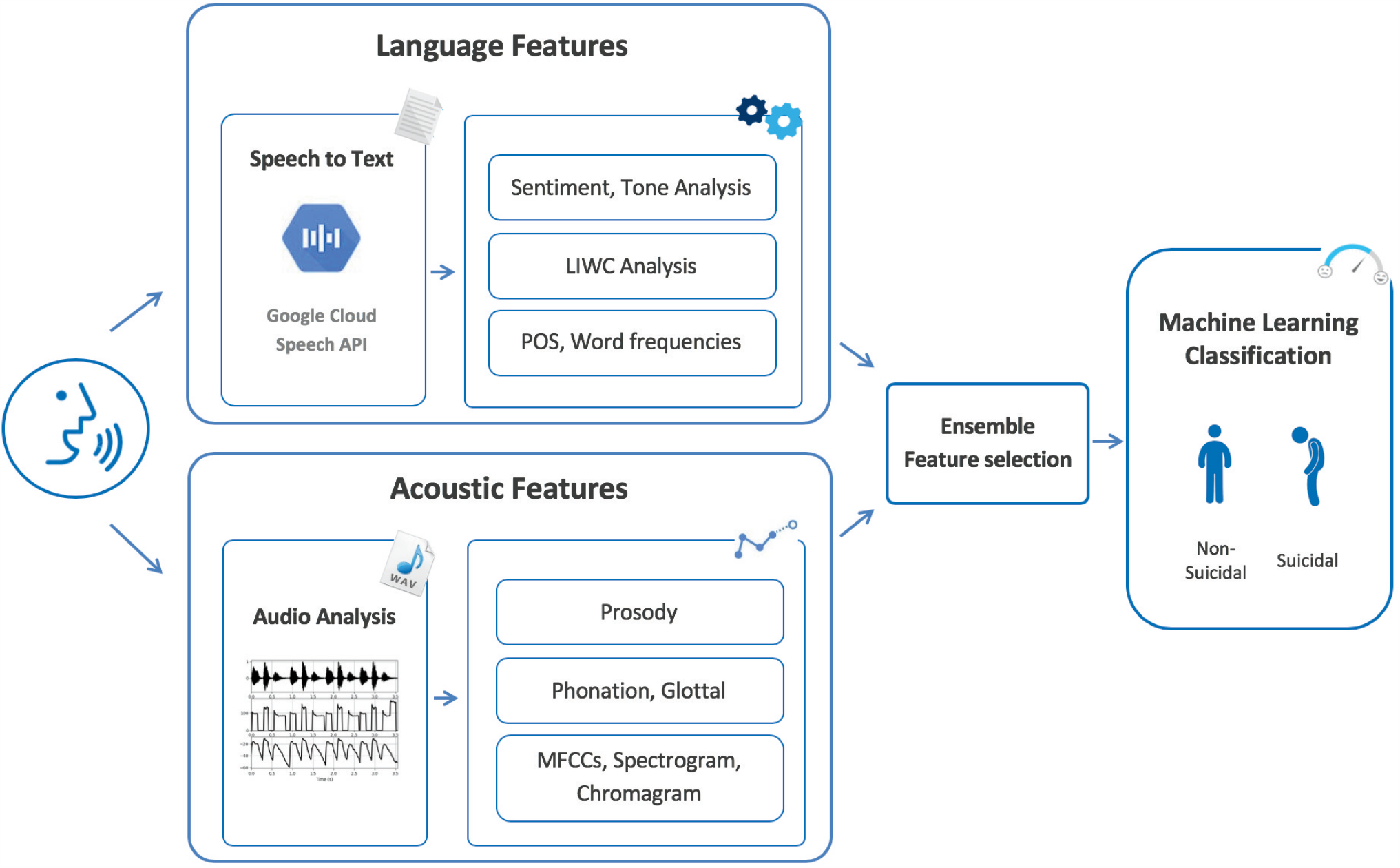
Outline of the study procedure. Acoustic features were extracted using pyAudioAnalysis and DisVoice audio python libraries. Audios were transcribed using Google Speech-to-Text API. Linguistic features were extracted using LIWC. POS and word frequency features were extracted using NLTK. Sentiment and tone analysis was performed using NLTK, Watson Tone Analyzer, Azure Text Analytics, and Google NLP.. We perform an ensemble feature selection to identify a subset of predictive features. We use different machine learning and deep learning techniques to build a suicidality classification model.

#### Acoustic Features

We extracted a total of 508 acoustic features from each of the 588 recordings. We used pyAudioAnalysis,^26^ an audio signal analysis python library, to extract short-term feature sequences using a frame size of 50 milliseconds and a frame step of 25 milliseconds (50% overlap). Then, we calculated recording level features as statistics on the short-term features (mean, maximum, minimum, median, standard deviation). The pyAudioAnalysis features include: zero crossing rate, energy and entropy of energy, chroma vector and deviation, spectral features composed of centroid, spread, entropy, flux, rolloff and Mel-Frequency Cepstral Coefficients (MFCC).

The second set of acoustic features were extracted using DisVoice,^27^ a python framework for feature extraction of pathological speech. We computed several prosodic features from continuous speech based on duration, fundamental frequency (F0), and energy. Phonation-based features were computed from sustained vowels and continuous speech utterances. For continuous speech, we computed the degree of unvoiced segments in addition to seven descriptors over voiced segments (first and second derivative of F0, jitter, shimmer, amplitude perturbation quotient, pitch perturbation quotient, logarithmic energy) then we derived higher-order statistics for each recording (mean, std, skewness, kurtosis). From sustained vowels, we computed 9 glottal features (variability of time between consecutive glottal closure instants (GCI), average and variability of opening quotient (OQ) for consecutive glottal cycles, average and variability of normalized amplitude quotient (NAQ) for consecutive glottal cycles, average and variability of H1H2: difference between the first two harmonics of the glottal flow signal, average and variability of Harmonic richness factor (HRF) and 4 statistics were derived (mean, std, skewness, kurtosis). All computed variables were then normalized to a range of zero to one.

#### Linguistic Features

All audio files were transcribed automatically using Google speech-to-text API, a speech recognition tool that achieves above 95% accuracy in speech recognition tasks.^28^ No quality checks were performed on the transcribed text corpus, as one of our hypotheses was to assess the feasibility of an automated approach of both acoustic and linguistic analysis of speech. Subsequently, we used the transcribed text and various Natural Language Processing (NLP) techniques to extract different sets of textual features.

##### Parts of Speech (POS)

We use the NLTK library^29^ to perform POS tagging on the text from each recording to generate features representing word classes and lexical categories. Furthermore, we compute word frequencies of absolutist terms which, in previous research, have been found to be associated with suicidal ideation.^30^

##### Sentiment Analysis

Given the psychological nature of suicide ideation, assessing the general polarity and emotions of the recordings is necessary. We compute sentiment scores and emotion level scores to detect joy, fear, sadness, anger, analytical, confident, and tentative tones in the language used by veterans. To perform the sentiment analysis we used the following tools and APIs: NLTK, IBM Watson Tone Analyzer, Azure Text Analytics, and Google NLP.

##### Linguistic Inquiry and Word Count program (LIWC)

The LIWC software^31^ is a computational text analysis tool that has been extensively used in the mental health space to explore various text corpora for hidden insights from linguistic patterns. We use the program to produce 94 features per recording, based on validated dictionaries covering a wide range of categories to assess different psychological, affective, and linguistic properties.

##### Text visualization

We further analyze the text using Scattertext,^32^ a text visualization tool to understand differences in speech between suicidal and non-suicidal veterans. The tool uses a scaled f-score, which takes into account the category-specific precision and term frequency. While a term may appear frequently in both groups, the scaled f-score determines if the term is more characteristic of one category versus another. We exclude stopwords (such as “the”, “a”, “an”, “in”) from the text corpus.

### Statistical analysis

We computed a total of 679 acoustic and linguistic features to understand speech in suicidal veterans. To compare suicidal and non-suicidal speech, we investigated these features by checking their statistical significance and magnitude of effect size. We used chi-square test for categorical variables and kruskal-wallis *H-*test for both continuous and ordinal variables. All raw *p*-values (*p*-raw) were adjusted for multiple testing using the Bonferroni correction where *p*-adj = *p-raw* x n, where n is the number of independent tests. We define statistical significance as *p-adj*<0.05. We also calculated the effect size using *epsilon*-squared (ϵ^*2*^) to understand how strong is the influence of a variable.^33,34^ The goal of this first analysis is to infer any significant relationships between the characteristics of speech and suicidality.

### Machine learning approach

The second analysis we performed on the extracted set of features is based on Machine Learning (ML). ML is an analytical approach that can uncover hidden and complex patterns to help generate actionable predictions in clinical settings.^35^ An essential step of any ML procedure is feature selection to reduce redundant variables and identify a stable subset of features. This can help create models that are easier to interpret and implement in real-life settings. We implemented an ensemble feature selection approach to select the top performing features across multiple selectors. This approach is known to improve the robustness of the selection process, especially in high-dimensional and low sample size.^36^ In particular, we used algorithms with built-in feature importance or coefficients such as ridge, lasso, random forest, and recursive feature elimination using logistic regression. For each algorithm, the best subset of features is selected and scores are assigned to each single feature. A mean-based aggregation is used to combine the results and provide a ranking of the top important and stable features.

In many clinical use cases, the outcome of interest is only represented by a few cases. We observed this class imbalance in our dataset with 1 suicidal recording for every 6 non-suicidal. To computationally deal with this imbalance, we used the SMOTE technique^37^ to oversample the minority class in the training sets after partitioning the data during the learning process. It is essential to oversample after data partitioning to keep the test data representative of the original distribution of the dataset and avoid information leakage that can lead to overly optimistic prediction results.^38^

We investigated different supervised classification algorithms on the selected features and evaluated the results. Specifically, we applied six algorithms: logistic regression (LR), random forest (RF), support vector machines (SVM), XGBoost (XGB), k-nearest neighbors (KNN), and deep neural network (DNN). Prediction performance was assessed using the area under the receiver operating characteristic curve (AUC), which indicates how capable a model is at distinguishing between classes. Although this metric can be optimistic for imbalanced datasets, it still shows a relative change with better performing models, especially when higher sensitivity is desired (i.e. detection of suicidal recordings is more important).^39^ Additionally, we report sensitivity, specificity, and accuracy. Since our data is imbalanced, it was important to assess the performance of the models based on all the metrics jointly. We used the Youden index^40^ to identify the optimized prediction threshold to balance sensitivity and specificity.

For model evaluation and selection, we performed a nested cross-validation (CV) learning approach where we split the data into a 5-fold inner and a 5-fold outer CV. During each iteration of the nested CV, we kept 1 outer fold for testing (20% of the samples) and used the 4 remaining folds in the 5-fold inner CV to search for the optimal model. We used a grid-search method in the inner loop to tune the different classification algorithms across a wide range of their respective hyperparameter settings. The final generalization error was estimated by averaging AUC scores over the outer test splits. We used nested CV, as opposed to regular k-fold CV, to reduce overfitting and produce stable and unbiased performance estimates that can generalize to unseen data.^41–43^

The data partitioning applied during the nested CV was stratified. This means that each fold of the CV split had the same class distribution as the original dataset (1:6 ratio). Further, given the longitudinal aspect of the dataset, multiple recordings can belong to the same participant and may have different suicidality labels across time. This potentially introduces data leakage where recordings from the same participant end up in both training and test folds. Since our goal was to build participant-independent models, we conducted a subject-wise CV to mirror the clinical use-case scenario of screening in newly recruited subjects.^44^

We built 3 different models to assess the predictive performance of acoustic and linguistic features separately and also when combined. The recordings were considered independent of the type of question asked or when they were recorded. In addition, we evaluated different minimum word counts and minimum audio length cutoffs for the inclusion of the recordings in the modeling.

## Results

### Demographics and recordings characteristics

Between May 2016 and January 2020, 149 veterans were recruited for a clinical intervention for Gulf War Illness. Of these, 25 Veterans didn’t submit any audio recordings. The remaining 124 participants submitted 588 recordings via the data collection mobile app. The average age of this group was 52.4 years (std= 9.4) and the majority of participants were male veterans (79%). Additional demographic characteristics are presented in **Table 1**.

**Table 1.** Background and characteristics of participants in the study, N=124

|  |  |  |
| --- | --- | --- |
| **Age, mean (std)** |  | 52.4 (9.4) |
| **Sex, n (%)** | Female | 26 (21.0) |
|  | Male | 98 (79.0) |
| **Race, n (%)** | Black or African-American | 86 (69.4) |
|  | Other | 12 (9.7) |
|  | White | 26 (21.0) |
| **Ethnicity, n (%)** | Hispanic or Latino | 5 (4.0) |
|  | Non Hispanic or Non Latino | 108 (87.1) |
|  | Unknown | 11 (8.9) |
| **Employment status, n (%)** | Employed Full-time | 47 (37.9) |
|  | Employed Part-time | 7 (5.6) |
|  | Other | 5 (4.0) |
|  | Receiving Disability Benefits | 38 (30.6) |
|  | Retired | 21 (16.9) |
|  | Unemployed | 6 (4.8) |
| **Education, n (%)** | Associate degree | 18 (14.5) |
|  | Bachelor degree | 35 (28.2) |
|  | High School diploma/GED | 25 (20.2) |
|  | Master's degree | 32 (25.8) |
|  | Other | 10 (8.1) |
|  | PhD/Doctorate degree | 4 (3.2) |
| **Partnership status, n (%)** | Divorced or separated | 31 (25.0) |
|  | Married or living with a partner | 83 (66.9) |
|  | Never married | 10 (8.1) |

Out of 588 audios, 504 were non-suicidal and 84 suicidal. All veterans recorded at least once, with a maximum of eight (21.7% of veterans). 74 veterans (59.6%) recorded at least 4 recordings. During week 0 and week 8, participants were asked to record two separate audios. After transcribing the audios, 15 recordings had no text transcriptions (5 suicidal and 10 non-suicidal). These audios were then manually verified and eventually excluded from the study, as they were either empty or had short intelligible speech.

### Acoustic Analysis

Average audio length was 44.19 seconds (std= 52.27). There were no significant differences between suicidal and non-suicidal recordings in audio length, loudness, or duration of pauses. Suicidal recordings were mainly different from non-suicidals in terms of energy. Suicidal speech had a lower standard deviation of energy contours for voiced segments *(p-adj <0.001*, ϵ^*2*^=*0.043)*, a lower kurtosis *(p-adj <0.001*, ϵ^*2*^=*0.06), and a* skewness closer to zero *(p-adj <0.001*, ϵ^*2*^=*0.052)* which reflect respectively flatter, less bursty, and less animated voice.

Suicidal speech had lower voiced tilt *(p-adj = 0.04*, ϵ^*2*^=*0.028)* and less energy entropy (*p-adj = 0.04*, ϵ^*2*^=*0.027*) thus displaying less vocal energy and less abrupt changes. Suicidal speech also exhibited a lower standard deviation of delta MFCC11 *(p-adj = 0.004*, ϵ^*2*^=*0.035)*, delta MFCC12 *(p-adj = 0.004*, ϵ^*2*^=*0.032)*, and delta MFCC1 *(p-adj = 0.05*, ϵ^*2*^=*0.023)*. The decrease in time derivatives (delta) of MFCC coefficients indicates a lack of variance of energy over time in suicidal speech which can be interpreted as dull and more monotonous voices. Additionally, suicidal veterans produced speech that was irregular in time and displayed high variability between consecutive GCIs (*p-adj = 0.035*, ϵ^*2*^=*0.028)*, which can be interpreted as breathier voices.

### Linguistic Analysis

Average word count was 70.96 words per recording (std= 93.76) with an average of 15.05 words per sentence *(std= 9.62)*. There were no significant differences between suicidal and non-suicidal recordings in word count or words per sentence. Suicidal participants used more possessive pronouns *(p-raw = 0.005*, ϵ^*2*^=*0.005*) and more superlative adverbs *(p-raw = 0.005*, ϵ^*2*^=*0.008)*. The analysis of the LIWC scaled scores showed that suicidal participants also used more family references (e.g. daughter, dad, aunt) *(p-raw = 0.014*, ϵ^*2*^=*0.010)* and more family male references (e.g. boy, his, dad) *(p-raw < 0.001*, ϵ^*2*^=*0.021)*. While, non-suicidal recordings contained more agentic language (e.g. win, success, better) *(p-raw = 0.035*, ϵ^*2*^=*0.007)*. There were no significant differences between the two groups in sentiment scores or usage of negative or positive emotion words. After adjusting for multiple testing, no linguistic features were significant.

Scattertext analysis (**Figure 2**) outlines the top words used by both suicidal and non-suicidal veterans. The tool analyzed over 40 thousand words from the text corpus to assign a scaled f-score to each word. Ranking words by f-score can help identify which terms are more characteristic of suicidality versus non-suicidality. Top words used by suicidal veterans were: “certainly”; “pills”; “real”; “knees”; “month”; “old”; “CPAP; “got”; “happened”; “stop”; “VA”; “certain”; “doctor”; “daily basis”. Top words used by non-suicidal veterans were: “energy”; “little bit”; “areas’; “‘aware”; “following”; “function”; “trying”; “find”; “noticed”; “bit”; “improve”; “days”; “group”; “meditation”.

**Figure 2:**
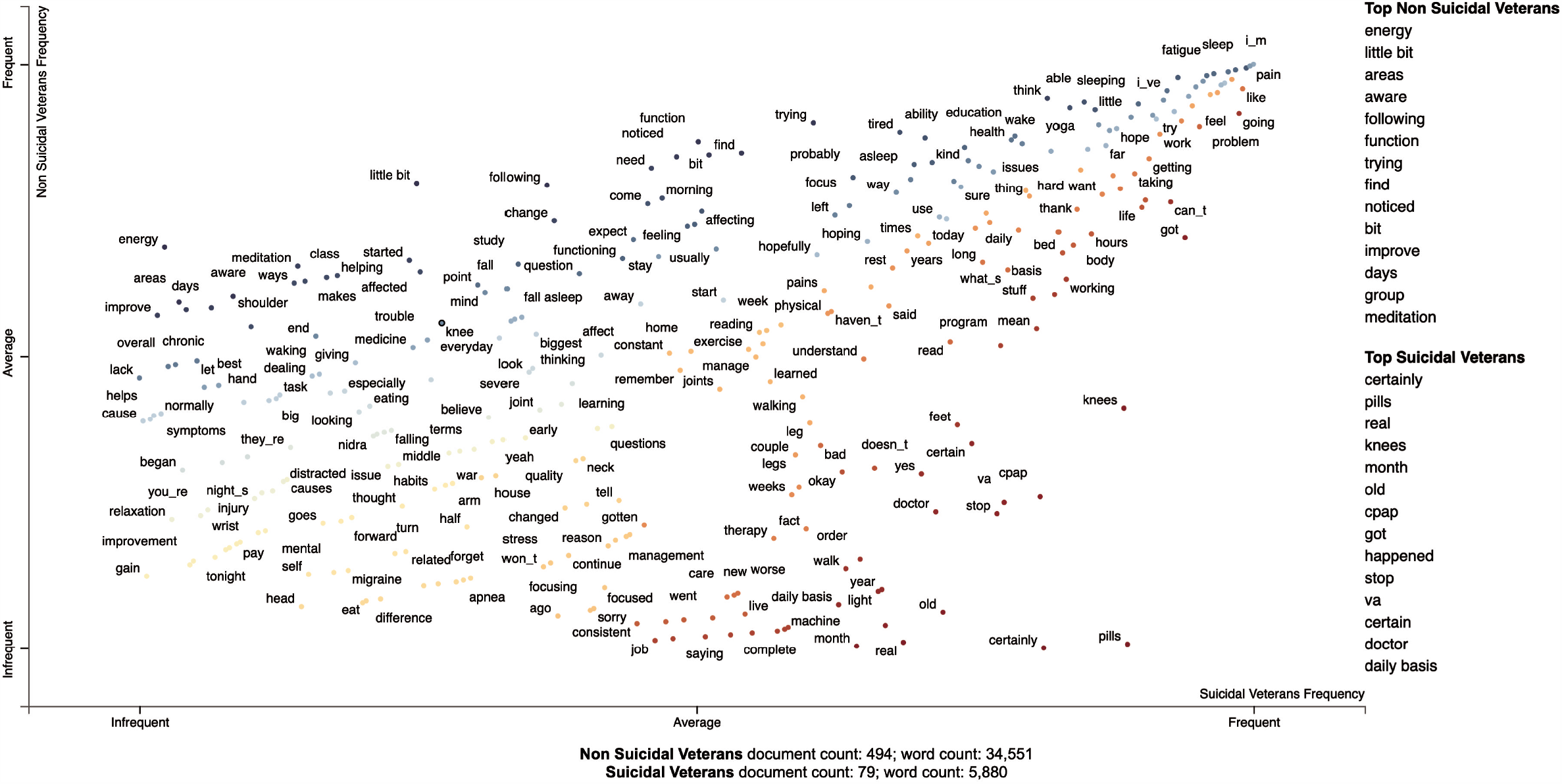
Scattertext visualization of words associated with both suicidal and non-suicidal groups. The red dots on the right lower side of the plot represent terms that are more associated with suicidal ideation compared to the blue dots which indicate terms more associated with non-suicidality.

### Selected features and prediction performance

**Table 2** presents the top 15 acoustic and linguistic features retained by the ensemble feature selection approach. These variables were used for the combined modeling (acoustic + linguistic) which yielded the best results. Out of the selected features, four were linguistic and assessed the use of superlative adverbs, possessive pronouns, personal nouns, and agentic language. The remaining features were acoustic and related to energy dynamics, MFCC, F0, and glottal flow measurements (i.e. OQ, NAQ).

**Table 2.** Description of the top 15 acoustic and linguistic features (rank-ordered by importance) retained for the combined machine learning modelling.

| Feature | Type | Description |
| --- | --- | --- |
| **Delta energy entropy (max)** | Acoustic (Prosody) | Entropy of energy can be interpreted as a measure of abrupt changes. |
| **Delta energy (mean)** | Acoustic (Prosody) | Dynamic changes in energy relate to the perceptual characteristics of pitch and loudness. |
| **Energy contour (kurtosis)** | Acoustic (Prosody) | The kurtosis of the energy contour for voiced segments. |
| **Delta F0 (std)** | Acoustic (Prosody) | First derivative of the fundamental Frequency. Reduced F0 can indicate low pitch and a flatter voice. |
| **MFCC5 (max)** | Acoustic (MFCC) | 5th MFCC coefficient. It can describe vocal tract changes in voice spectral energy. |
| **Superlative adverbs** | Linguistic (POS) | Use of superlative adverbs (e.g. biggest, hardest..) |
| **OQ (skewness)** | Acoustic (Glottal) | OQ is a measurement of the glottal flow. It can differentiate between a breathy and tense voice. |
| **Achievement language** | Linguistic (LIWC) | Use of agentic language as defined by the LIWC achievement dictionary (words such as win, success, better...) |
| **Delta chroma 2 (min)** | Acoustic (Prosody) | A representation of spectral energy. Chroma-based features are also referred to as "pitch class profiles". |
| **Proper nouns** | Linguistic (POS) | Use of singular proper nouns. |
| **Delta energy (median)** | Acoustic (Prosody) | Dynamic changes in energy relate to the perceptual characteristics of pitch and loudness. |
| **Delta chroma 1 (median)** | Acoustic (Prosody) | A representation of spectral energy. Chroma-based features are also referred to as "pitch class profiles". |
| **Delta MFCC5 (max)** | Acoustic (MFCC) | The frame-based delta of the 5th MFCC coefficient. It can measure vocal tract changes. |
| **NAQ (mean)** | Acoustic (Glottal) | Average NAQ is a measure of the glottal flow that can differentiate between a breathy and tense voice. |
| **Possessive pronouns** | Linguistic (POS) | Use of possessive pronouns (e.g. my, his, hers..) |

We evaluated different word count (WC) cutoffs to identify the minimum utterances needed to better discriminate suicidality in recordings. Classification performances increased as we increased the WC minimum. The best classification results were obtained for recordings with a minimum of 25 words. **Table 3** presents the performance of the classifiers.

**Table 3.** Classification results for suicidal ideation based on acoustic and linguistic features.

| Feature set | Model | Specificity (std) | Sensitivity (std) | Accuracy (std) | AUC (std) |
| --- | --- | --- | --- | --- | --- |
| **Acoustic** | **KNN** | 0.52 (0.23) | 0.78 (0.13) | 0.55 (0.19) | 0.69 (0.11) |
|  | **LR** | 0.64 (0.15) | 0.78 (0.11) | 0.66 (0.13) | 0.78 (0.12) |
|  | **RF** | 0.64 (0.12) | 0.76 (0.17) | 0.65 (0.09) | 0.76 (0.06) |
|  | **SVM** | 0.54 (0.30) | 0.74 (0.18) | 0.56 (0.25) | 0.63 (0.25) |
|  | **XGB** | 0.74 (0.21) | 0.67 (0.18) | 0.73 (0.17) | 0.77 (0.08) |
|  | **DNN** | 0.70 (0.06) | 0.66 (0.20) | 0.70 (0.05) | 0.75 (0.06) |
| **Linguistic** | **KNN** | 0.69 (0.15) | 0.38 (0.14) | 0.65 (0.11) | 0.52 (0.07) |
|  | **LR** | 0.57 (0.24) | 0.66 (0.20) | 0.58 (0.18) | 0.62 (0.06) |
|  | **RF** | 0.64 (0.17) | 0.76 (0.09) | 0.65 (0.14) | 0.74 (0.07) |
|  | **SVM** | 0.63 (0.33) | 0.48 (0.40) | 0.61 (0.24) | 0.49 (0.15) |
|  | **XGB** | 0.70 (0.16) | 0.67 (0.17) | 0.69 (0.13) | 0.72 (0.04) |
|  | **DNN** | 0.50 (0.15) | 0.66 (0.25) | 0.52 (0.11) | 0.63 (0.14) |
| **Acoustic and Linguistic** | **KNN** | 0.61 (0.22) | 0.78 (0.22) | 0.63 (0.19) | 0.69 (0.15) |
|  | **LR** | 0.74 (0.14) | 0.78 (0.19) | 0.75 (0.11) | 0.77 (0.12) |
|  | **RF** | 0.70 (0.16) | 0.84 (0.09) | 0.72 (0.13) | 0.80 (0.06) |
|  | **SVM** | 0.86 (0.13) | 0.52 (0.32) | 0.82 (0.09) | 0.64 (0.27) |
|  | **XGB** | 0.79 (0.11) | 0.74 (0.18) | 0.78 (0.08) | 0.77 (0.05) |
|  | **DNN** | 0.68 (0.06) | 0.70 (0.15) | 0.68 (0.04) | 0.77 (0.08) |

The XGB classifier overall performed best on acoustic features with a sensitivity of 0.67 (std=0.18), specificity of 0.74 (std=0.21), accuracy of 0.73 (std=0.17), and an overall AUC of 0.77 (std=0.08). The LR classifier performed slightly better in terms of AUC with 0.78 (std=0.12) and sensitivity of 0.78 (std=0.11) but had a lower specificity of 0.64 (0.15) which resulted in a much lower accuracy of 0.66 (0.13).

On linguistic features, RF performed better than other classifiers overall with a sensitivity of 0.76 (std=0.09), specificity=0.64 (std=0.17), accuracy of 0.65 (std=0.14), and an overall AUC of 0.74 (std=0.07). XGB classifier performed better in terms of accuracy 0.69 (std=0.13) and specificity of 0.70 (std=0.16) but performed lower on sensitivity and AUC.

As shown in **Table 3**, combining both acoustic and linguistic features improved the models. RF classifier correctly identified suicide ideation in veterans with an overall sensitivity of 0.84 (std=0.09), specificity of 0.70 (std=0.16), accuracy of 0.72 (std=0.13), and an AUC of 0.80 (std=0.06). Overall, tree-based models (RF and XGB) performed best on this dataset.

## Discussion

In the present study, we conducted a two-part analysis. First, we investigated the importance of the extracted acoustic and linguistic features using statistical significance. Second, we evaluated an ensemble feature selection approach to identify a subset of features that can be used in an ML model to detect suicidality in veterans. We demonstrated that characteristics of speech can be useful in differentiating between suicidal and non-suicidal recordings. Our findings also indicate that audios collected outside the clinical setting, using a mobile app, can be used to classify suicidality with an overall AUC of 0.80.

In the first analysis, we sought to understand characteristics of suicidal speech in veterans and infer significant relationships. A notable finding of the study are the 3 features related to energy contour of voiced segments (std, kurtosis, skewness). These variables displayed the largest effect size and indicated the strongest difference between the suicidal and non-suicidal groups. We found that suicidal veterans spoke in voices that were flatter, less bursty, and less animated. Additional energy-based variables such as speech tilt, energy entropy, and MFCC coefficients, indicated speech in suicidal veterans that had less vocal energy, less abrupt changes, and was more monotonous. The analysis of the glottal flow parameters related to GCIs indicated a more breathy voice quality in suicidal veterans. These findings are in line with results from previous studies on other risk groups. For example, a study that examined GCIs, OQ, and NAQ, found that suicidal adolescents had a more breathy voice quality compared to non-suicidal adolescents.^18^ In addition, multiple research studies used levels of energy dynamics and MFCC features to distinguish controls and depressed subjects who subsequently attempted suicide.^12,17,45^ The general dullness of speech and reduction in energy has also been correlated with PTSD in veterans compared to controls.^14^

The linguistic analysis produced no significant variables. Nevertheless, we observed trends indicating more superlative adverbs, possessive pronouns, and personal nouns in suicidal speech. On the other hand, non-suicidal veterans used more agentic language based on the LIWC achievement score. The scatter text analysis, although exploratory in nature, provided frequently used words among suicidal and non-suicidal veterans. Overall, we found that suicidal veterans spoke with certainty (e.g. certain, certainly…) discussing topics such as chronic pain (pills, knees) or sleep problems (CPAP machine) when describing their general health in the past weeks and months. Conversely, non-suicidal veterans used action verbs and words indicating improvements (e.g. function, improve, trying, find, noticed, aware…). Interestingly, chronic pain and apnea discussed by suicidal veterans have been both linked to suicide as risk factors ^46,47^. In addition, previous research on internet forums showed that suicidal subjects use more possessive pronouns and more absolutist words.^30,48^

Building a classification model for suicidality was the second part of the analyses presented here. The results show that acoustic-based models performed better (AUC=0.78) than models based on linguistic features alone (AUC=0.74). Given the links between suicidality and language, we also explored advanced NLP techniques to improve the linguistic models, such as word and document embeddings. Classification using embeddings provided weak results (not presented), which was mainly due to the relatively small text corpus. Such techniques can be promising for the classification of suicidality if applied to a much larger corpus. A key finding of the study is that we achieved higher accuracies by combining both acoustic and linguistic features (AUC= 0.80). This is in line with previous research on depression and other mental states where fusion of different modalities such as audio, text, and visuals helped improve prediction results.^13,49–51^ Accuracies reached by our models are comparable to previous research on suicidality and speech in other risk groups, however, the few published studies either relied on smaller sample size or didn’t discuss what important features went into their final models. ^12,18,20,52,53^

This is the first study to assess suicidality in US veterans using speech. It is also, to the best of our knowledge, the first study on suicidality to collect recordings longitudinally from participants in a real-life setting using a mobile app. This is essential since previous work on suicidality and speech used structured clinical interviews which, although can provide high quality voice corpora, might also introduce interviewer- and potentially environment-induced biases.^9^ Collecting data digitally without the involvement of another human can reduce the stress associated with the fear of being judged and hence produce less biased recordings. Additionally, subjects using a mobile app might be willing to disclose more.^54,55^ Such an approach has the potential to be fully automated and implemented for longitudinal and context-aware monitoring by collecting audio diaries from veterans at high risk.

The impact of findings from this study may be limited by a number of factors. We relied on self-report to indicate whether a subject was suicidal or not at the time of the recording. Hence, it is possible that some of the recordings were mislabeled if a participant was not willing to divulge their suicidal state. Further, audios ranged from a few seconds to several minutes long and were made in a variety of everyday life settings which could have introduced background noise and quality issues. An additional limitation may stem from possible confounders given that participants might suffer from other mental states such as depression and anxiety. This makes it difficult to determine whether the identified features are solely linked to suicidality or might be linked to other comorbid mental states that are more likely to present in suicidal subjects. Future work assessing other mental states along with suicidal ideation could help improve the classifiers and further validate the identified features. Improvements to the classifiers could also come from different fusion methods of acoustic and linguistic features such as ensemble modeling or from context-based analysis where the questions asked are also weighted in the models.

## Conclusions

We showed that speech analysis is a promising approach for detecting suicidal ideation in veterans. We also demonstrated that recordings collected longitudinally outside the clinical setting, using a mobile app, can be utilized for such analysis. Using both statistical and predictive modeling, we identified a set of important acoustic and linguistic markers of speech that can be useful in classifying suicidality in these recordings. The choice of the ML approach and dimensionality reduction techniques were important to optimize the performance of the classifiers and provide realistic estimations on unseen data. Further external validation and optimization will be needed to validate and improve these findings. Overall, our work supports the feasibility of an automated approach of both acoustic and linguistic analysis of speech in everyday life settings, which holds the promise for real-time suicidality assessment in high-risk veterans.

## Data Availability

Public sharing of the data is not possible.

## Acknowledgements

This project has been funded partly by Georgetown-Howard Universities Center for Clinical and Translational Science (GHUCCTS) (UL1-TR001409) and partly with Federal funds (5 I01 CX000801 02) from the U.S. Department of Veterans Affairs Medical Center. The content of this work does not necessarily reflect the policies or position of the US Government.

